# PERIPHERAL ARTERIOVENOUS MALFORMATIONS AND THEIR RESPONSE TO TREATMENT MODALITIES: A SINGLE SURGEON’S EXPERIENCE AT TERTIARY CARE HOSPITALS

**DOI:** 10.1101/2021.09.15.21262513

**Authors:** Rashid Usman, Duaa Ajaz Hussain, Muhammad Jamil Malik, Muhammad Waseem Anwar, Kishwar Ali

## Abstract

**Background:** Peripheral arteriovenous malformations (AVMs) are congenital defects resulting in abnormal connections between veins and arteries. We investigated a group of patients with peripheral AVMs to determine whether there were any gender differences in disease presentation and the response of AVMs to various modalities of treatment.

**Methodology:** The patients in this cross-sectional study were divided into two groups based on gender and their demographic data, clinical presentation at the time of diagnosis and response to treatment was assessed. Both surgical and nonsurgical treatment options were used. Nonsurgical options employed in the study included immunotherapy, embolotherapy and ultrasound-guided foam sclerotherapy (UGFS)

**Results:** Out of 43 patients, 74.4% were females with a male to female ratio of 1:3. The mean age at presentation in males was 27 years and 17 years in females. 60% of the male patients presented with high-flow AVMs while 81% of the female patients presented with low-flow lesions instead. Half of the AVMs in males were on the trunk whereas, in females, 93.9% were on the extremities. UGFS alone was used in 95.3% of patients while 32.5% of patients underwent UGFS followed by surgical excision. One patient was treated with sirolimus. In 4.6% of cases, embolization followed by surgical excision was performed. Recurrence was recorded in 20.9% of cases.

**Conclusions:** The clinical presentation of AVMs is notably different among the two genders. Sclerotherapy and embolotherapy proved to be effective treatment options. Larger studies, however, are needed to substantiate these claims.

## INTRODUCTION

AVMs are congenital defects arising due to errors in embryogenesis that result in abnormal connections between veins and arteries. Treatment of AVMs can broadly be classified as surgical and nonsurgical [1]. Nonsurgical options include sclerotherapy, embolotherapy and immunotherapy. AVMs are rare, accounting for only 10-15% of all clinically significant CVMs [2] with an estimated prevalence of 5-613 in 100,000 [3].

There is a paucity of data regarding peripheral AVMs, especially of studies conducted locally in Pakistan. Our research intends to serve as a pilot study for this purpose. We investigated a small group of patients with peripheral AVMs to explore whether there were any gender differences in disease presentation and the response of AVMs to various modalities of treatment.

## MATERIALS & METHODS

This retrospective, multicentre cross-sectional study was conducted at three locations: Combined Military Hospital Lahore, Midcity Hospital Lahore and Combined Military Hospital Rawalpindi. Approval was obtained from the Institutional Review Board (IRB) of Combined Military Hospital Rawalpindi (approval number 68/18/05/21). All patients diagnosed with an AVM that had been treated from January 2016 to June 2020 were considered potentially eligible for the study. Informed written consent was obtained from all patients. Patients that declined consent or had a documented anaphylactic reaction to the sclerosant were excluded. Pregnant patients and children under five years were also not included. Lastly, patients lost to a minimum of six months follow-up were also excluded. After applying the above criteria, we reached a sample size of 43 patients.

Ultrasound-guided foam sclerotherapy (UGFS) where applicable, was performed using a 30 mg/ml injection of Sodium Tetradecyl Sulfate (STS). Tessari’s method was used to mix STS, water and air in a 1:2:2 ratio. Post-sclerotherapy compression was maintained for two weeks. Sirolimus was administered as a twice-daily oral dose of 0.5mg/m^2^ for 12 cycles over a year. This was followed by three sessions of UGFS as described.

Monthly follow-ups were performed for at least six months. Ultrasound imaging was performed with GE Logiq Book (General Electric, Massachusetts, United States). Clinical examination and ultrasound Doppler were performed at each follow-up to assess treatment efficacy. The treatment modality and total number of sessions was recorded. Recurrence was defined as the re-appearance of an AVM after Doppler-recorded healing. The recurrence rate and other complications were recorded.

Statistical analysis was performed using Statistical Packages for the Social Sciences program (SPSS) version 24 (IBM Inc., Armonk, USA). Quantitative variables like age were expressed as mean ± standard deviation. Qualitative variables like gender were expressed as frequencies and percentages. Patients were divided into two groups based on gender and were compared using Chi-square Test. A p-value less than or equal to 0.05 was considered statistically significant.

## RESULTS

### Patient and AVM characteristics

Among 43 cases, there were 10 (23.2%) males and 32 (74.4%) females with a male to female ratio of 1:3. Mean age at time of presentation in males was 27± 6 years (range 19 -40 years) and 17± 4 years (range 11-22 years) in females. The difference in age between the groups was statistically significant as the p-value calculated was 0.003. In terms of AVM type, in males, 60% of AVMs were of the high-flow type while in females, the low-flow type of AVMs was more prevalent (81%). Moreover, 50% of AVMs in males were on the trunk followed by head and neck (30%) and limbs (20%). In females, this ratio was reversed with the limbs being the most prevalent site (93.9%) and the trunk being the least affected site (3%). These findings are further clarified in Table 1.

**Table 1:**
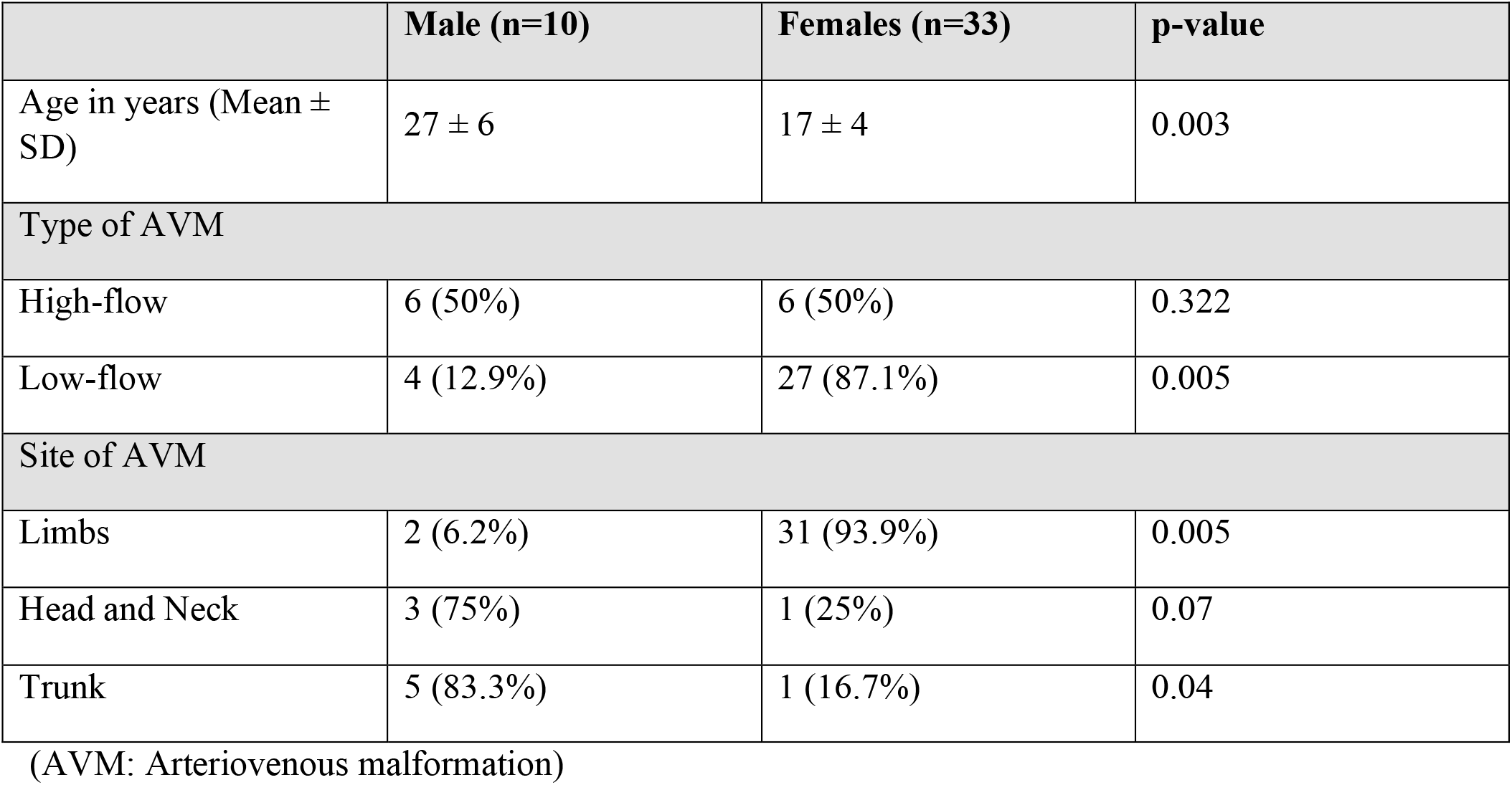
Characteristics of male and female patients at time of presentation

|  | Male (n=10) | Females (n=33) | p-value |
| --- | --- | --- | --- |
| Age in years (Mean ± SD) | 27 ± 6 | 17 ± 4 | 0.003 |
| Type of AVM |  |  |  |
| High-flow | 6 (50%) | 6 (50%) | 0.322 |
| Low-flow | 4 (12.9%) | 27 (87.1%) | 0.005 |
| Site of AVM |  |  |  |
| Limbs | 2 (6.2%) | 31 (93.9%) | 0.005 |
| Head and Neck | 3 (75%) | 1 (25%) | 0.07 |
| Trunk | 5 (83.3%) | 1 (16.7%) | 0.04 |

(AVM: Arteriovenous malformation)

### Treatment modalities and response

41 (95.3%) patients were treated with UGFS, out of which 26 received UGFS as the sole mode of treatment. All 26 patients had low-flow AVMs. Within this group, 14 (53.8%) patients were cured after a single session of UGFS, while 8 (30.7%) and 4 (15.3%) patients needed two and more than two sessions respectively for complete healing. A total of 14 (32.5%) patients underwent UGFS followed by surgical excision. 9 (64.2%) of these patients had high flow AVMs while 5 (35.7%) had low flow AVMs. All patients with high-flow AVMs required more than one session of UGFS.

One patient had a huge high-flow AVM unamenable to formal excision. He was treated with 12 cycles of oral sirolimus followed by three sessions of UGFS. On follow up, persistent small pockets of active blood flow were present. Nevertheless, a significant reduction in the lesion’s size was observed.

Preoperative embolization followed by surgical excision was performed in 2 cases (4.6%). Care was taken to perform the surgery within the same week as the embolization.

### Recurrence and Complications

Recurrence was recorded in 9 (20.9%) patients, 6 (66.6%) of which had high-flow AVMs. Therefore, the recurrence rate was 50% for high-flow AVMs and 9.6% for low-flow AVMs.

In patients who underwent UGFS, local pain and swelling were noted in 21 (51.2%) cases. Skin necrosis and ulceration occurred in eight (19.5%) patients, while superficial local thrombophlebitis was observed in seven (17%) patients. However, these complications resolved with conservative management. Other complications included hematoma and seroma formation in 12 (27.9%) and 10 (23.2%) patients respectively, along with the development of a superficial surgical site infection in 3 (6.9%) patients.

## DISCUSSION

AVMs are congenital vascular lesions that may not be apparent until later in life. These lesions require angiography to reach a diagnosis [1]. AVMs are treated early because they can be life or limb-threatening. A large AVM can cause high-output cardiac failure due to the absence of an intervening capillary bed. Artery steal syndromes can also occur due to the presence of AVMs, resulting in underperfusion of tissues. Moreover, a large AVM closely related to a long bone can result in limb-length discrepancy either by limb undergrowth or overgrowth [4].

The male to female incidence of AVMs in our study was 1:3 which contrasts with the commonly reported 1:1 ratio for AVMs [5]. Furthermore, the average age at presentation in female patients was younger compared to male patients. Two factors might account for these findings. First, AVMs tend to grow under hormonal influences, enlarging after puberty and due to pregnancy [6]. Therefore, hormonal exposure could act as a confounding factor, leading to an earlier pickup in women. Secondly, our study offers a snapshot view of the disease which might result in over-representation of female cases as these become clinically apparent earlier while an equal number of male patients may have been picked up over a longer course of time.

Male patients tended to present with an AVM involving the trunk whereas the limbs were more frequently involved in female patients. While there is some data to suggest an effect of sex on the presentation of intracranial AVMs, more studies are needed before similar conclusions can be drawn for peripheral AVMs [7].

Another difference we found was the nature of AVMs. Males more often presented with high-flow AVMs while females commonly presented with low-flow AVMs. This can be accounted for if we consider the presence of “little AVMs”, as defined by Stein et al. as malformations on the spectrum between AVMs and venous malformations [8]. These lesions present clinically as AVMs but behave radiologically as low-flow lesions. Although the predominant opinion is that there are no gender differences in the prevalence of low-flow malformations, one series did, however, find a female preponderance, similar to the findings in our study [9].

Endovascular techniques including sclerotherapy and embolotherapy were initially introduced as a second line of treatment for lesions not amenable to formal excision, but are now often combined with surgery due to the higher success rate of this approach [10]. Sclerotherapy has long been used to treat vascular malformations. The sclerosant we used was STS (sodium tetradecyl sulfate), which has detergent properties that injure the vessel [11], resulting in fibrosis and shrinkage [12] of the lesions. Adverse effects of STS include swelling and necrosis, which were observed in more than half of our patients that underwent UGFS. Another adverse effect is hemoglobinuria, however, there were no cases in our study that developed this complication.

STS is an effective option for the treatment of AVM, as demonstrated by eventual complete resolution in the 26 patients in our study who underwent UGFS as monotherapy. Other studies treating AVMs with STS have had similar results [13,14].

A wide variety of embolics may be used for embolotherapy, from mechanical coils to liquid embolics[15]. The agent we used was NBCA (N-butyl cyanoacrylate), a glue that polymerizes when it comes in contact with blood, forming an instant cast of the vessel [16]. Preoperative embolization reduces flow through the AVM, resulting in reduced blood loss during surgery [17]. The most significant complication that can arise after embolization is the occlusion of non-target vessels, which can result in end-organ ischemia. Awareness of inciting factors such as injection technique is important to minimize harm, as NBCA glue deposition is ultimately unpredictable in vivo [18].

Sirolimus (rapamycin) is an mTOR inhibitor used to treat vascular malformations. mTOR (mammalian target of rapamycin) is an important regulator of cellular proliferation [19] and its inhibition can prevent angiogenesis [20].Treatment with sirolimus did not result in resolution in our study, as evident by persistent flow in the AVM. This is in line with the findings of other studies which showed none or only minor improvements at best with the use of sirolimus for AVMs [21,22]. However, it should be considered as a palliative treatment option for management of symptoms.

## LIMITATIONS

The most important limitation of our study is that the sample size was restricted by both the rarity of the disease and the multiple exclusion criteria employed. This is justified as we aimed to run a pilot study to explore whether any notable differences were present in the patient groups studied. However, despite the level of significance found, these findings cannot be generalized to the overall population unless larger, more powerful studies are run to further validate these claims. These findings may further be influenced by the potential presence of a healthcare access bias. The number of patients treated for AVMs therefore, is an underestimate, as not all patients have access to the imaging facilities required to reach a diagnosis and subsequent referral for specialist intervention.

## CONCLUSION

Based on the findings of this cross-sectional study, there are notable gender differences in the clinical presentation of AVMs. Male patients with these lesions tend to present much later in life than female patients. Moreover, AVMs show variance in site and type based on gender. Male patients were more likely to present with high-flow AVMs in the trunk region whereas female patients typically presented with low-flow AVMs on the distal extremities. AVMs planned for surgical treatment should first undergo sclerotherapy or embolotherapy to reduce perioperative complications and the recurrence rate. Palliative care, on the other hand, can be provided via novel options such as immunotherapy with sirolimus.

Given the lack of similar data on peripheral AVMs in the Pakistani population, our research intended to serve as a pilot study. These findings were therefore only exploratory in nature. Future longitudinal studies are needed before any decisive claims about these relations can be made. Larger studies are also needed to make recommendations for treatment regimens at the level of the general population.

## Data Availability

The authors confirm that the data supporting the findings of this study are available within the article [and/or] its supplementary materials.

## REFERENCES

1. Mulligan P, Prajapati H, Martin L, Patel T. Vascular anomalies: classification, imaging characteristics and implications for interventional radiology treatment approaches. The British Journal of Radiology. 2014;87(1035):20130392.

2. Lee B, Lardeo J, Neville R. Arterio-venous malformation: how much do we know?. Phlebology: The Journal of Venous Disease. 2009;24(5):193–200.

3. Stapf C, Mohr J, Pile-Spellman J, Solomon R, Sacco R, Connolly E. Epidemiology and natural history of arteriovenous malformations. Neurosurgical Focus. 2001;11(5):1–5.

4. Bertino F, Braithwaite K, Hawkins C, Gill A, Briones M, Swerdlin R et al. Congenital Limb Overgrowth Syndromes Associated with Vascular Anomalies. RadioGraphics. 2019;39(2):491–515.

5. Lee B, Baumgartner I, Berlien H, Bianchini G, Burrows P, Do Y et al. Consensus Document of the International Union of Angiology (IUA)-2013. Current concept on the management of arterio-venous management. Int Angiol [Internet]. 2013 [cited 17 June 2021];32(1):9–36. Available from: https://pubmed.ncbi.nlm.nih.gov/23435389/

6. Manzocchi Besson S, Jastrow Meyer N, Bounameaux H, La Scala G, Calza A, Yilmaz H et al. Multiple arteriovenous malformations caused by RASA1 gene mutation presenting during pregnancy – a case report and review of the literature. Vasa. 2019;48(3):276–280.

7. Tong X, Wu J, Lin F, Cao Y, Zhao Y, Ning B et al. The Effect of Age, Sex, and Lesion Location on Initial Presentation in Patients with Brain Arteriovenous Malformations. World Neurosurgery. 2016;87:598–606.

8. Stein M, Guilfoyle R, Courtemanche D, Moss W, Bucevska M, Arneja J. The “Little AVM”. Plastic and Reconstructive Surgery Global Open. 2014;2(7):e187.

9. Nosher J. Vascular anomalies: A pictorial review of nomenclature, diagnosis and treatment. World Journal of Radiology. 2014;6(9):677.

10. Sohail M, Bashir M, Ansari H, Khan F, Assumame N, Awan N et al. Outcome of Management of Vascular Malformations of Lip. Journal of Craniofacial Surgery. 2016;27(6):e520–e524.

11. Stuart S, Barnacle A, Smith G, Pitt M, Roebuck D. Neuropathy after Sodium Tetradecyl Sulfate Sclerotherapy of Venous Malformations in Children. Radiology. 2015;274(3):897–905.

12. Ahmad S. Efficacy of Percutaneous Sclerotherapy in Low Flow Venous Malformations - A Single Center Series. Neurointervention. 2019;14(1):53–60.

13. Kirkpatrick D, Frenette A, Hasham H, Custer B, Lemons S, Collins Z et al. Successful Percutaneous Treatment of an Arteriovenous Malformation of the Toe. Annals of Vascular Surgery. 2020;65:288.e5–288.e8.

14. Sitra G, Sivasankari T, Vishwanath R, Kayalvizhi E. A new venture with sclerotherapy in an oral vascular lesion. Journal of Basic and Clinical Pharmacy. 2015;6(1):40.

15. Potts M, Zumofen D, Raz E, Nelson P, Riina H. Curing arteriovenous malformations using embolization. Neurosurgical Focus. 2014;37(3):E19.

16. Khurana A, Hangge P, Albadawi H, Knuttinen M, Alzubaidi S, Naidu S et al. The Use of Transarterial Approaches in Peripheral Arteriovenous Malformations (AVMs). Journal of Clinical Medicine. 2018;7(5):109.

17. Kansy K, Bodem J, Engel M, Freudlsperger C, Möhlenbruch M, Herweh C et al. Interdisciplinary treatment algorithm for facial high-flow arteriovenous malformations, and review of the literature. Journal of Cranio-Maxillofacial Surgery. 2018;46(5):765–772.

18. Hill H, Beecham Chick J, Hage A, Srinivasa R. N-butyl cyanoacrylate embolotherapy: techniques, complications, and management. Diagnostic and Interventional Radiology. 2018;.

19. Sehgal S. Sirolimus: its discovery, biological properties, and mechanism of action. Transplantation Proceedings. 2003;35(3):S7–S14.

20. Adams D, Trenor C, Hammill A, Vinks A, Patel M, Chaudry G et al. Efficacy and Safety of Sirolimus in the Treatment of Complicated Vascular Anomalies. Pediatrics. 2016;137(2):e20153257.

21. Gabeff R, Boccara O, Soupre V, Lorette G, Bodemer C, Herbreteau D et al. Efficacy and Tolerance of Sirolimus (Rapamycin) for Extracranial Arteriovenous Malformations in Children and Adults. Acta Dermato Venereologica. 2019;:0.

22. Freixo C, Ferreira V, Martins J, Almeida R, Caldeira D, Rosa M et al. Efficacy and safety of sirolimus in the treatment of vascular anomalies: A systematic review. Journal of Vascular Surgery. 2020;71(1):318–327.

